# Targeted Free Swimming for Women, Girls, and Older Adults in Tower Hamlets: A RE-AIM Evaluation Study Protocol

**DOI:** 10.64898/2026.10.05.26361900

**Authors:** Kristen Clements, Yvanna Todorova, Aku Makawa, Perfect E. Asamoah, Paul Lester, Denis M. Ngina, Lauren Sherar

## Abstract

**Background:** Residents of the London Borough of Tower Hamlets experience higher levels of inactivity than national averages, with particularly low participation among women, girls, older adults, and people living in deprived communities. In response, Tower Hamlets launched a targeted free-swimming programme in July 2024, providing free swimming sessions, lessons, coaching clinics, and planned childcare support across four leisure centres. The programme aims to reduce barriers to physical activity, increase participation among underserved groups, and promote health equity. This evaluation seeks to assess the programme’s reach, implementation, acceptability, effectiveness, and sustainability.

**Methods:** A mixed-methods evaluation guided by the RE-AIM framework will assess the reach, implementation, acceptability, and effectiveness of the Tower Hamlets free-swimming programme. Quantitative analysis of routine registration and attendance data will examine participant characteristics, programme uptake, and engagement patterns. Qualitative interviews and focus groups with programme users, non-users, and delivery staff will explore experiences, barriers, facilitators, and perceived impacts. Quantitative and qualitative findings will be triangulated to provide a comprehensive understanding of programme delivery and outcomes.

**Expected Outcome:** The evaluation is expected to generate evidence on the extent to which the programme engages priority populations, including women, girls, and older men. It is anticipated that the study will identify factors influencing programme uptake and sustained engagement, including cultural appropriateness, accessibility, and implementation processes. The evaluation will also provide insights into perceived impacts on physical activity, wellbeing, social connectedness, swimming confidence, and health inequalities. Furthermore, the study is expected to identify the programme components most valued by participants, including free access, women-only sessions, swimming lessons, coaching opportunities, and childcare provision.

**Implication policy and practice:** The findings will provide actionable evidence to support local authority decision-making regarding the refinement, continuation, and potential expansion of the free-swimming programme. The study will contribute to the evidence base on place-based physical activity interventions designed to address health inequalities in diverse urban populations. For policymakers, the evaluation will offer insights into how subsidised leisure services can improve access to physical activity, enhance community wellbeing, and promote equitable participation among underserved groups.

## 1.0 Background and Rationale

### 1.1 Geographical and Socioeconomic Context of Tower Hamlets

Tower Hamlets, one of London’s oldest boroughs, was established in 1965 and is situated on the north bank of the River Thames, immediately east of the City of London (LBTH Research, 2009). The borough is governed by Tower Hamlets London Borough Council and had a population of 310,000 in 2021, reflecting a 22% increase from the 255,000 recorded in 2011 (Eade, 2024). This rapid growth makes it one of the fastest growing and most densely populated boroughs in England.

The borough is among the most ethnically and culturally diverse in London. According to the 2021 Census, 44.4% of residents are Asian, Asian British, or Asian Welsh (up from 41.1% in 2011), 39.4% White (down from 45.2%), 7.3% Black, Black British, Black Welsh, Caribbean, or African, 5.0% Mixed or Multiple ethnic groups, and 3.9% Other ethnic groups. Within this, British Bangladeshis form the largest ethnic group, accounting for around 34.6% of residents (Corporate Strategy and Improvement Team, 2024). This diversity shapes engagement with health and recreational activities, as cultural factors such as modesty and gender-specific preferences influence participation in activities like swimming. The borough reflects considerable diversity in gender and sexuality, with 7.2% of adults identifying as Lesbian, Gay, Bisexual, or Other LGBTQ+ groups, and around 1% having a gender identity differing from the sex assigned at birth - (Corporate Strategy and Improvement Team, 2024).

In addition, socioeconomic challenges in the borough are significant. A 2017 joint study by Trust for London and the New Policy Institute ranked Tower Hamlets as the second most deprived London borough, based on indicators such as child poverty, unemployment, and income inequality (Trust for London, 2025). The borough exhibits stark contrasts in wealth, with a small proportion of very high earners alongside a large share of households on low incomes, leaving nearly half of residents reliant on benefits. Benefit claims increased sharply during the COVID-19 pandemic, from around 9,100 in May 2019 to over 21,000 at the height of the pandemic, settling at approximately 13,800 in 2021, nearly 5,000 above pre-pandemic levels (Tatari & Teymuroglu, 2023).

Social conditions further compound these challenges. Overcrowding, where households have more occupants than can be reasonably accommodated within available rooms affects 17% of residents, one of the highest rates in London, while 11% experience fuel poverty, leaving many vulnerable to rising energy costs and the wider cost-of-living crisis (Green, 2017).

Overall, 31% of residents live in England’s most deprived areas, placing Tower Hamlets 57th out of 317 local authorities nationwide. This deprivation disproportionately impacts children and older adults, contributing to increased risks of physical inactivity and associated poor health outcomes (Tower Hamlets Public Health Department, 2016).

Together, these demographic, social, and economic factors translate directly into significant public health challenges, setting the stage for high prevalence of both physical and mental health conditions. The combination of rapid population growth, ethnic diversity, socioeconomic inequality, and limited access to resources highlights the urgent need for targeted, inclusive public health interventions to improve health and reduce inequalities in the borough.

### 1.2 Evidence and Gaps Underpinning the Targeted Free-Swimming Programme

Physical inactivity remains one of the most pressing public health challenges globally, contributing to avoidable deaths, chronic disease, and rising healthcare costs. Globally, inactivity is estimated to cost around £27 billion each year through healthcare expenditure, productivity losses, and years of healthy life lost (Shilton et al., 2024). In the UK, physical inactivity is linked to approximately one in six deaths and imposes an annual economic burden of £7.4 billion, including nearly £1 billion in direct NHS costs (Public Health England, 2016). These figures reflect not only the scale of the problem but also its persistence. Compared with the 1960s, the UK population is now around 20% less active, and if current trends continue, activity levels could fall by a further 35% by 2030, an alarming trajectory with implications for both population health and the sustainability of the health system.

Recent national data reinforce the urgency of reversing this decline. Between November 2023 and November 2024, nearly one in four adults in England (22%) were classified as physically inactive, engaging in fewer than 30 minutes of moderate activity each week. In contrast, around two-thirds (67%) met the Chief Medical Officers’ recommended 150 minutes of weekly moderate-intensity activity, while fewer than half (47.8%) of children and young people achieved the guideline of at least 60 minutes of daily physical activity (Sport England, 2025). These figures point to a systemic gap in opportunities, environments, and motivation to engage in regular movement across the life course. Addressing this gap requires localised and culturally relevant strategies capable of engaging communities who face the greatest barriers to participation.

In Tower Hamlets, the consequences of these national trends are especially evident. The borough experiences one of the highest burdens of non-communicable diseases in London, driven in part by persistently low levels of physical activity. Cardiovascular disease mortality stands at 112 per 100,000, substantially above the England average of 78 while the prevalence of type 2 diabetes is projected to rise from 15,612 diagnosed cases in 2016 to nearly 25,000 by 2030 (Chowdhury & Huda, 2016). These conditions account for a significant proportion of local morbidity and healthcare utilisation, mirroring national economic losses of £7.4 billion annually for cardiovascular disease and £14 billion for diabetes (McKay et al., 2014). The link between inactivity and health inequality is particularly pronounced in Tower Hamlets, where deprivation, ethnicity, gender, and age intersect to limit access to active lifestyles (Quin et al., 2022).

Consequently, promoting physical activity is central to improving health outcomes and reducing inequalities in the borough. Swimming presents a particularly effective and inclusive means of doing so. As a low-impact, whole-body activity, swimming can be sustained across age groups and physical abilities, including for those with joint or mobility limitations. Importantly, it can be adapted to meet diverse cultural needs such as through gender-specific sessions that encourage women and girls’ participation (Staley et al., 2024). Evidence consistently supports the health benefits of swimming: a large-scale survey of over 80,000 participants found that regular swimming reduces the risk of premature death by 28% and cardiovascular mortality by 47% (Swim England, 2017). Moreover, swimming supports mental wellbeing, with nearly half a million adults in the UK using it to manage mental health conditions; of these, 43% report reductions in anxiety and depression (Swim England, 2017). Among children, swimming fosters physical fitness, confidence, and social connection, while helping to address rising levels of obesity (Massey et al., 2022).

Beyond its physical and mental health benefits, swimming holds particular relevance for Tower Hamlets given the borough’s dense urban environment and limited access to affordable, inclusive recreational spaces. Public pools offer one of the few accessible environments where residents of all ages, abilities, and backgrounds can engage in safe and structured physical activity. In this context, swimming represents not only a means of improving fitness and wellbeing but also of fostering social connection and community cohesion. Yet, participation remains uneven, only around half of residents from ethnically diverse communities can swim 25 metres, compared with 86% of white communities, and females of all ethnicities are less likely than males to meet this benchmark (Tower Hamlets Public Health Department, 2022). These disparities illustrate broader inequalities in access, confidence, and opportunity, underscoring the need for interventions that make swimming both culturally acceptable and practically accessible across the borough’s diverse population.

Taken together, the evidence positions swimming as a practical, culturally adaptable, and evidence-based intervention capable of addressing the intertwined challenges of physical inactivity, chronic disease, and inequality. In a borough such as Tower Hamlets, marked by high health need, social diversity, and unequal access to active environments, targeted free-swimming initiatives provide a compelling, locally relevant strategy for promoting sustained participation in physical activity and improving health equity. The next section introduces the Targeted Free-Swimming Programme developed in response to these challenges.

### 1.3 Overview of targeted Free-Swimming Programme

The Be Well initiative launched a targeted free-swimming programme (FSP) on 15 July 2024. The FSP in Tower Hamlets is designed to increase physical activity among groups traditionally underrepresented in exercise, such as women and girls aged 16 and over, and all male residents aged 55 and over. Eligible participants register online and receive a free membership card, granting access to designated either ‘swim for fitness’ or ‘women-only’ sessions across four leisure centres in Tower Hamlets namely Mile End Park Leisure Centre and Stadium, Poplar Baths Leisure Centre and Gym, Tiller Leisure Centre, and York Hall Leisure Centre. These centres offer tailored sessions to accommodate the diverse needs of the community.

The FSP in Tower Hamlets is being implemented in three phases. Phase 1 commenced in July 2024 and includes the core free swimming activity across four leisure centre pools, providing up to 2,805 free sessions per week, including 186 Women Only sessions. This represents approximately 25% of the total “Swim for Fitness” capacity, with 5% reserved for walk-in participants who may have difficulty booking online. Since its inception, the programme has seen substantial participation, with over 20,000 residents registered by July 2025. Notably, women and girls aged 16 and over account for more than 18,000 of these participants, while nearly 3,900 residents aged 55 and over are also actively engaging in the sessions. This represents a significant increase in female participation, with women and girls now comprising 82% of users, up from just over 40% before the scheme launched (Tower Hamlets Council, 2025).

Participation spans a broad range of age groups, demonstrating the programme’s wide community appeal. Younger adults have engaged strongly, with over 3,400 members aged 16–24 and more than 5,700 aged 25–34. An additional 4,448 residents aged 35–44 and over 3,100 aged 45–54 are also actively participating (Tower Hamlets Council, 2025). The introduction of women-only sessions has been particularly impactful in addressing cultural and gender-specific barriers, fostering a more inclusive environment. Together, these elements highlight the programme’s success in increasing access, promoting equitable participation, and supporting Tower Hamlets’ broader public health goals.

Phase 2 began in September 2024, focusing on targeted teaching and coaching to encourage new swimmers and build confidence in the water. The programme aimed to support up to 200 participants each year through 12-week courses delivered as part of the existing swim teaching provision at each pool. Each course was designed to accommodate eight non-swimmers per pool and ran four times a year, providing regular opportunities for new participants to get involved. Additional swim clinics offered tailored support for individuals with basic swimming ability or limited confidence, helping them develop the skills needed to swim safely and independently. By combining structured lessons with accessible coaching, Phase 2 sought to increase participation, improve swimming proficiency, and strengthen the wider FSP by engaging residents who might otherwise face barriers to pool access.

Phase 3 is designed to introduce free or subsidised creche facilities, either permanent or “pop-up” arrangements at the leisure centres, to encourage greater participation from women with children in the programme. At present, the service is exploring options for providing these facilities alongside additional Women Only sessions. While the precise level of demand is not yet known, potential spaces include the former café area at Poplar, the soft play area or meeting rooms at Mile End Stadium and Tiller Leisure Centre, though York Hall has more limited space. Furthermore, the programme is considering flexible “pop-up” creches delivered by external childcare providers, subject to regulatory requirements, to better understand demand and the impact on uptake and inequalities. This phase aims to engage at least 10% of new users who have not previously accessed council swimming pools and contribute to the council’s target of increasing female participation in leisure centre activities by 5%.

The FSP also seeks to strengthen workforce diversity and community participation by training 20 new female lifeguards and enrolling 200 adults in the Learn to Swim programme. These efforts complement the 723 weekly free-swimming sessions already in place, including Women Only sessions that enhance accessibility and inclusion.

### 1.4 Why the need for evaluation?

After a year of implementing the FSP as part of the Be Well initiative, the London Borough of Tower Hamlets Council’s Public Health Division has commissioned an independent evaluation to understand how the programme is contributing to residents’ health and general wellbeing. The evaluation is considered timely, providing an opportunity to assess who the programme is reaching, how effectively it is engaging underrepresented groups, and the extent to which it is achieving its intended objectives. It will also explore barriers and facilitators to participation, variation in delivery across sites, and the perceived value of the programme among participants and staff.

Findings from this evaluation will generate critical insights to inform future programme refinement, delivery, and investment decisions. In particular, the evidence will help the Council strengthen accessibility and engagement among populations who are eligible but less likely to participate, while enhancing the overall impact of the FSP on health, wellbeing, and equity outcomes in Tower Hamlets.

## 2.0 Working in Partnership

### 2.1 Co-production of the proposal

Partnership working is a core principle of the PHIRST initiative and is embedded throughout this evaluation. The evaluation was co-designed by the PHIRST Light research team in collaboration with the London Borough of Tower Hamlets (LBTH) and local partners, ensuring that the perspectives of those involved in implementing and delivering the FSP were incorporated from the outset.

In line with this collaborative approach, the PHIRST team worked closely with LBTH to identify key stakeholders connected to the programme at different stages of delivery. These stakeholders were invited to participate in a series of online co-production workshops held via Microsoft Teams, which focused on stakeholder mapping, the development of a logic model, and the design of the evaluation framework. Participants included representatives from Be Well staff, Tower Hamlets Council for Voluntary Service (THCVS), leisure centre managers, and members of the Public Health and Research teams within the local authority, alongside researchers from the PHIRST Light team and members of the Public Advisory Group (PAG).

The workshops were designed to build consensus on the central research aims and identify the most suitable methodologies for addressing them. Through these sessions, participants collaboratively developed and refined a draft programme logic model (see Appendix 1) and jointly agreed on the key components to be tested through the evaluation. A preliminary set of research questions was also proposed to guide discussion and ensure that stakeholder priorities were meaningfully reflected in the study design. The final evaluation questions were refined through iterative feedback and now represent a shared understanding of what constitutes relevant, practical, and impactful learning for local delivery.

Stakeholder engagement remains an ongoing feature of the evaluation. Partners will continue to contribute through co-production and dissemination workshops, helping to interpret findings, contextualise emerging insights, and ensure that the evaluation process remains inclusive, transparent, and grounded in local realities.

### 2.2 Public contribution

Public involvement is a core component of the PHIRST Light research team, with a strong focus on building capacity for inclusive and meaningful engagement within local authorities. In line with the National Institute for Health and Care Research (NIHR) principles on patient and public involvement (PPI), the team has established a central PAG made up of members of the public and local service users.

The PAG oversees all PPI activities and is co-led by our PPI coordinator, Pam Rees, and academic lead, Prof. Jo Morling. In addition to this overarching group, we have supported the LBTH team in forming a project-specific PPI group to guide the evaluation design and delivery. This group includes six to eight individuals with lived experience of the FSP, and is coordinated by two PAG members, Cecily Henry and Marcia Hudson.

The group is actively involved in co-producing key elements of the evaluation, including recruitment strategies and participant materials, interview guides and dissemination plans.

These activities are primarily delivered through both online and face to face workshops and document reviews. All members are reimbursed for their time in line with NIHR guidance on honorarium payments. PPI contributors have been involved from the outset, participating in proposal development, stakeholder workshops, and research meetings. Their feedback has directly shaped the evaluation strategy and methodology, ensuring that lived experience remains central to the project.

## 3.0 Key evaluation aims, objectives and research questions

### 3.1 Research Questions

- How is the Free-Swimming Programme being implemented across Tower Hamlets, and what factors enable or constrain continuous delivery, access, and uptake?
- How do participants experience the Free-Swimming Programme, and what perceived impacts does participation have on their health and wellbeing?
- Who is accessing the Free-Swimming Programme, and to what extent does participation reflect the diversity and priority populations of Tower Hamlets?

### 3.2 Evaluation aim

To assess how the Tower Hamlets Free Swimming Programme is implemented and experienced in practice, and how delivery influences service reach, accessibility, and participants perceived health and wellbeing benefits

### 3.3 Evaluation objectives

To examine how the Free-Swimming Programme is delivered across Tower Hamlets and identify the factors that enable or constrain effective implementation, access, and uptake. (Implementation and Adoption)

To assess who is accessing the Free-Swimming Programme and the extent to which participation reflects the diversity and priority populations of Tower Hamlets

To explore how participants experience the Free-Swimming Programme and the perceived impacts of participation on their health and wellbeing

## 4.0 Methodological Overview

### 4.1 Study design

Informed by the logic model co-developed with Tower Hamlets Local Authority and local evaluation public contributors, this study will undertake a mixed methods process and outcome evaluation of the Tower Hamlets FSP. Framed within the RE-AIM framework (Reach, Effectiveness, Acceptability, Implementation, and Maintenance), the evaluation will examine both the contextual factors influencing implementation in real-world settings and preliminary programme outcomes.

The quantitative outcome evaluation will use service usage data to assess programme reach, participation trends, subgroup differences, and short-term effectiveness. Complementing this, the qualitative process evaluation will explore participant and stakeholder perspectives, focusing on barriers, facilitators, contextual influences, and perceived impacts on wellbeing and social connectedness.

Mixed-methods evaluations have been successfully applied in similar public health interventions, such as the National Exercise Referral Scheme in the UK, Walking for Health programmes, and school-based physical activity initiatives, where quantitative data captured measurable outcomes and qualitative insights informed implementation improvements and sustainability strategies. Applying this approach to the Tower Hamlets programme will generate actionable evidence to guide ongoing implementation, adaptation, and long-term sustainability, ensuring the programme effectively meets the needs of the community.

### 4.2 RE-AIM Framework: Application in this project

The RE-AIM framework provides a structured approach to examining how health interventions function in real-world settings across five interconnected dimensions: Reach, Effectiveness, Adoption, Implementation, and Maintenance. In this evaluation, the framework is adapted to reflect the nature of available data and the study’s focus on implementation and user experience.

- **Reach** refers to the number and characteristics of individuals who participate in the programme, enabling assessment of who the programme is engaging and the extent to which it reflects the diversity of the Tower Hamlets population.
- **Effectiveness (Perceived Outcomes)** focuses on participants’ self-reported experiences and perceived impacts of the programme on their health, wellbeing, and behaviours, recognising that causal outcome measurement is beyond the scope of this evaluation.
- **Adoption and Acceptability** considers both the uptake of the programme across delivery settings and how it is experienced by participants and providers. This includes perceptions of accessibility, cultural relevance, and value, as well as factors influencing engagement and participation.
- **Implementation** examines how the programme is delivered in practice, including variation across sites, adaptations to delivery, and contextual factors that influence access, uptake, and consistency.
- **Maintenance** explores the extent to which engagement with the programme is sustained over time and the factors that support continued participation, particularly among priority groups.

### 4.3 Data Collection Plans and Procedures

This study will draw on both quantitative and qualitative data to conduct the process and outcome evaluation.

#### 4.3.1 Quantitative data

Quantitative data in the form of routinely collected longitudinal usage data, are being gathered by the Tower Hamlets Local Authority. These data capture demographic information such as age, gender, ethnicity, postcode, parenting status etc alongside usage patterns of residents participating in the FSP across all four leisure centres. The data are stored within the Local Authority’s leisure management system, Gladstone.

In addition to demographic characteristics, the dataset is expected to include a range of usage indicators that can provide insights into participation trends and engagement over time. These may include the frequency of visits per user, duration of participation across months, recency of use based on the time since the last recorded visit, repeat participation rates indicating the proportion of returning users, the number of new registrations within defined periods, and centre-level utilisation patterns showing overall attendance across the four leisure centres. Together, these indicators will allow for an assessment of engagement trends, participation consistency, and the distribution of programme use across different demographic groups and settings.

Approximately **17,000 residents of Tower Hamlets** have registered for the FSP across the four participating leisure centres. This extensive dataset provides a valuable basis for examining programme reach and engagement patterns over time, as well as informing the sampling strategy for the qualitative components of the study.

#### 4.3.2 Qualitative data

Qualitative data will be collected to explore perceptions, experiences, and contextual factors influencing the implementation and outcomes of the free-swimming programme. The qualitative strand will involve in-depth interviews and focus group discussions with three key participant groups: service users, non-service users, and service providers or stakeholders. Data collection will aim to understand how the programme is perceived, accessed, and delivered within the Tower Hamlets community, as well as its perceived value, acceptability, and potential impact on wellbeing, physical activity, and social connectedness.

Interviews will be semi-structured, lasting approximately 30–45 minutes, and will be conducted either via videoconferencing platforms such as Microsoft Teams, by telephone, or in person, depending on participant preference. This flexible approach is designed to accommodate participants’ schedules and accessibility needs, acknowledging known challenges in reaching some segments of the community. With participants’ consent, interviews will be audio recorded and subsequently transcribed verbatim for analysis. Interviews will take place at locations convenient for participants, including their homes, workplaces, or other community settings.

All qualitative data will be collected by trained members of the research team with expertise in community-based qualitative research. The research team will also maintain reflective field notes to support contextual understanding during analysis. Data collection will continue until thematic saturation is reached across participant groups.

##### 4.3.2.1 Service Users

Service users include individuals who have accessed the FSP in the last 12 months, as well as those who regularly swim at participating leisure centres through paid memberships. Paid members will be eligible only if they have used the swimming facilities at least five times in the past six months. This ensures that all participants have enough recent experience to comment meaningfully on the programme and on how the introduction of the FSP may influence swimming behaviour, perceptions of fairness, and engagement with leisure centre services.

Discussions will explore participants’ awareness, accessibility, and attitudes towards the FSP. A core component will examine perceptions of the programme’s effectiveness in supporting health, wellbeing, and sustained engagement in physical activity. In addition to capturing the experiences of FSP users, the discussions will also engage paid membership swimmers to understand how the introduction of the FSP may have influenced their swimming habits, perceptions of value, and overall satisfaction with leisure centre services. Participants will be encouraged to reflect on their experiences across different leisure sites and service options. For example, comparing free sessions with structured swimming lessons and to identify perceived barriers and facilitators to participation. Finally, the conversations will invite reflections on both the positive and negative impacts of the programme on swimming confidence, physical activity levels, wellbeing, and sense of social connection.

##### 4.3.2.2 Non-service users

The second participant group will comprise non-service users, defined as individuals who are eligible to participate in the FSP but have not done so. Data from this group will be collected through two to three focus group discussions (FGDs), each involving approximately 6–8 participants. The use of focus groups is intended to encourage open dialogue and shared reflections, allowing participants to build on one another’s ideas and reveal collective perceptions that might not emerge in individual interviews.

These discussions will explore a range of topics, including reasons for non-participation, perceived barriers to swimming or accessing the service, and the cultural and social acceptability of the programme within the Tower Hamlets community. Attention will also be given to understanding how factors such as gender norms, religious considerations, affordability perceptions, awareness levels, and accessibility of facilities influence participation decisions.

Focus groups will be conducted either online or in person, depending on participant preferences, logistical feasibility, and accessibility needs. This flexible approach is expected to enhance participation from diverse community members and capture a broad spectrum of views, particularly from groups who may be underrepresented in programme uptake.

##### 4.3.2.3 Service providers

The third group will include service providers and key stakeholders, with the final sample size guided by the stakeholder mapping exercise. This group will include representatives from the Tower Hamlets Local Authority (public health and leisure departments), leisure centre staff, swim coaches, and others involved in the conception, design, delivery, and maintenance of the FSP. Interviews will examine programme implementation processes, resource and staffing implications, observed participant outcomes, contextual factors shaping delivery, partnership working, and opportunities for improvement, sustainability, and scale-up.

To ensure the evaluation is inclusive and accessible, we have incorporated several measures to meet the diverse needs of participants. Data collection tools and supporting documents have been redesigned to maximise legibility, including adaptations for an older adult with visual impairment and for individuals with dyslexia (e.g., increased font size, clear formatting, and reduced visual complexity). We also provide hard-copy materials for those who find on-screen reading or writing difficult.

We have engaged a public contributor to support the translation of research materials from English into Bangla, helping ensure cultural and linguistic relevance for participants from Bangladeshi communities. All translated materials will be reviewed and verified by a Research Assistant with relevant language expertise at the University of Nottingham to ensure accuracy and clarity.

In addition, we have built in time flexibility and financial support for participants with caring responsibilities to reduce barriers to attending interviews or workshops. These considerations are embedded across the evaluation to support meaningful participation and minimise disadvantage for all contributors.

### 4.4 Sampling Strategy and Participant recruitment

#### 4.4.1 Sampling Strategy and Size

A combination of convenience and snowball sampling will be used to recruit participants for both the in-depth interviews (IDIs) and FGDs. Participants will be identified through leisure centre staff and local networks to ensure accessibility and engagement from individuals who uses and does not use the FSP. However, we acknowledge that convenience-based recruitment can introduce selection bias, particularly if participants are drawn disproportionately from specific demographic or cultural groups, or from a single leisure centre. To minimise this risk and enhance representativeness, convenience and snowball sampling will be applied within a light quota–stratified framework. Quotas will be set to ensure diversity across leisure centres and, where possible, ethnic backgrounds, reflecting the demographic composition of Tower Hamlets and the varying patterns of FSP participation across sites.

For service users, we anticipate conducting approximately 20 IDIs, although the final number of interviews will be guided by data saturation, that is the point at which no new themes emerge. This sample size is expected to capture a diverse range of experiences, motivations, and perspectives among different user groups, including variations in age, gender, swimming ability, and frequency of use, thereby providing a comprehensive understanding of how the programme is perceived and experienced.

For non-service users, it is anticipated that three to four focus group discussions will be conducted with approximately six to eight participants in each group, yielding an estimated total of around 32 participants for this category.

The third participant group, comprising service providers will purposively be identified by the research team and PAG/PPI members. They will include at least two frontline staff members from each participating leisure centre, as well as intermediary or liaison staff who operate between management and delivery levels (e.g., data analysts, leisure commercial managers, or programme coordinators). In addition, three to five senior managers responsible for strategic decision-making and overall programme governance will be invited to participate. Overall, this group is expected to include approximately 10–15 participants.

#### 4.4.2 Participant recruitment

Eligible participants will be identified through stakeholder mapping and the local authority’s leisure service platform to contact individuals who previously consented to be contacted, with leisure service providers and swimming instructors assisting in distributing the study information sheet and invitation to eligible individuals via email or phone. Those who express interest will be contacted directly to confirm eligibility and discuss participation details, including their preferred mode of interview (online or in person at a designated meeting room within the leisure centre). Participants will also be invited to indicate their preferred interview times and any accessibility or comfort needs. Scheduling preferences will be collected securely via Calendly or through direct email communication, depending on participant preference.

Inclusion Criteria

- Resident of Tower Hamlets (at least a year)
- Women and girls aged 16 years and over, or men aged 55 years and over
- Aged 18 years or above at the time of participation

Exclusion Criteria

- Not a resident of Tower Hamlets
- Men younger than 55 years
- Individuals under 18 years of age

### 4.5 Overview of data collection plans

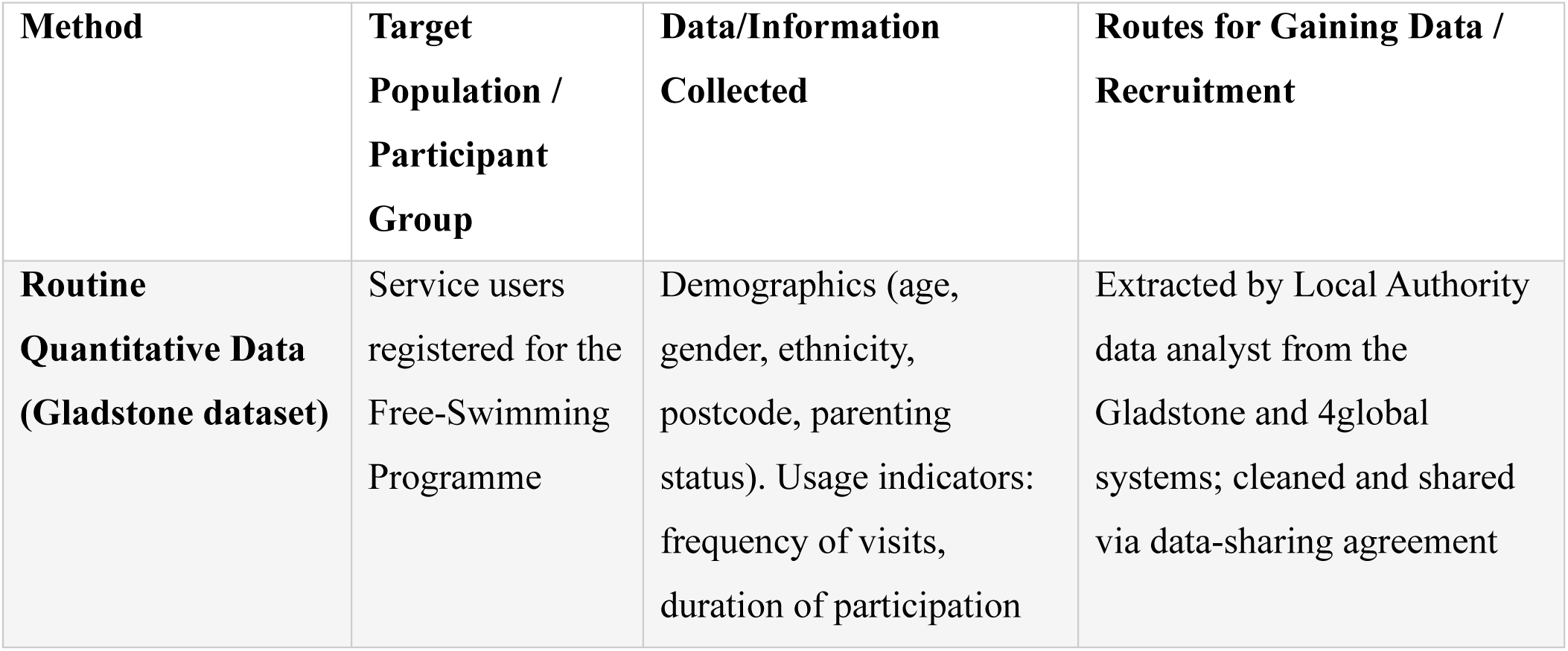
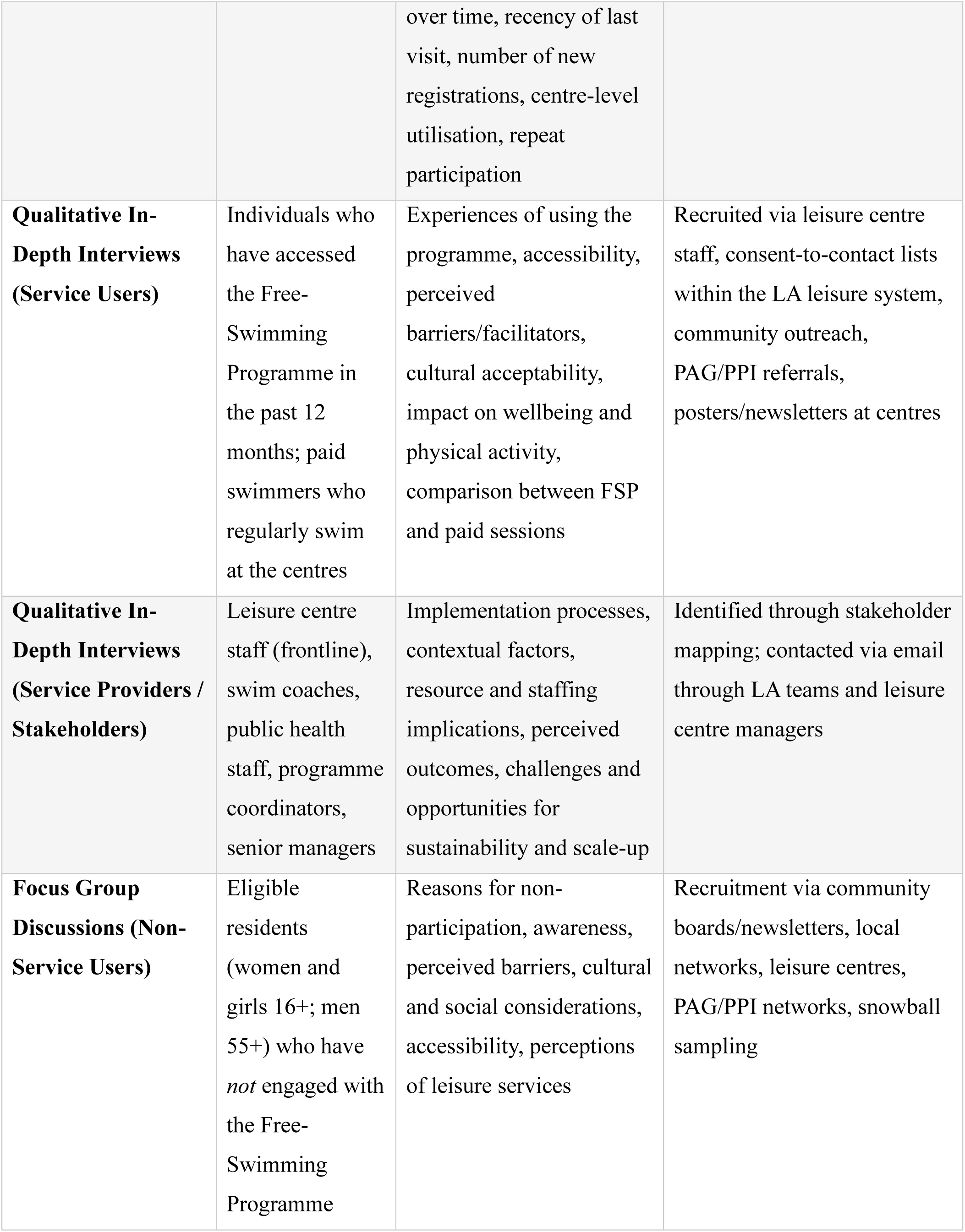
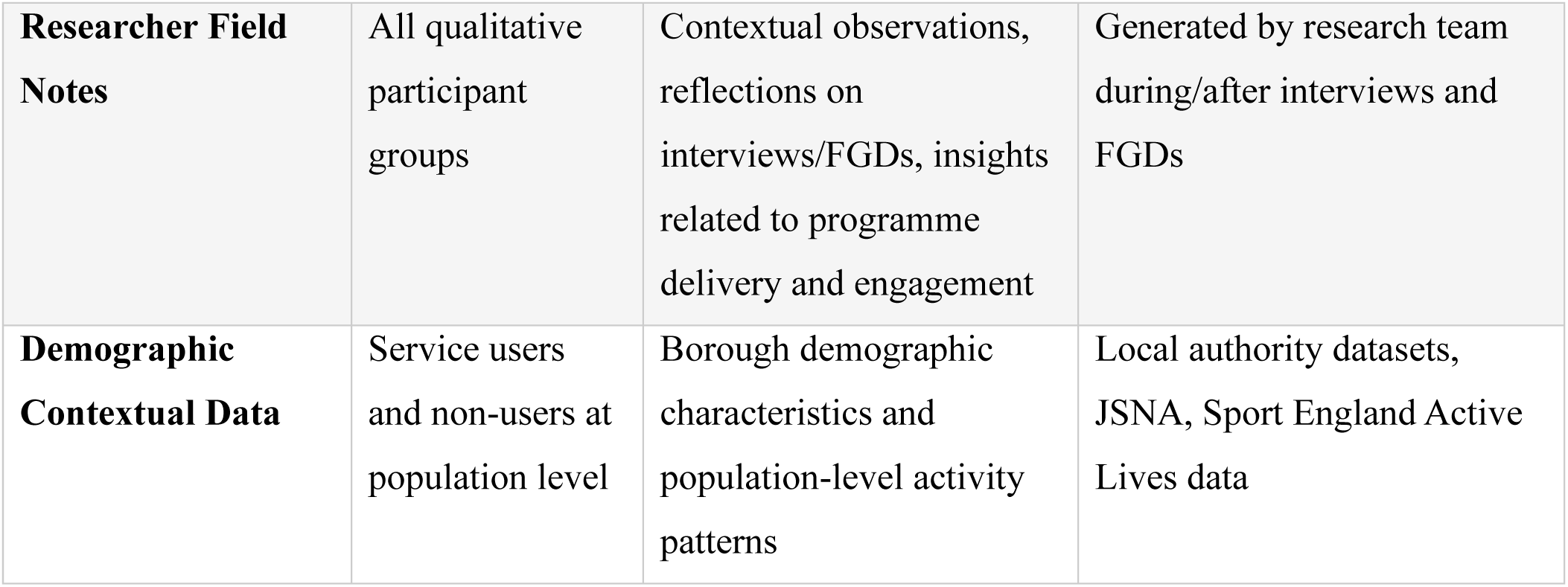

| Method | Target Population / Participant Group | Data/Information Collected | Routes for Gaining Data / Recruitment |
| --- | --- | --- | --- |
| **Routine Quantitative Data (Gladstone dataset)** | Service users registered for the Free-Swimming Programme | Demographics (age, gender, ethnicity, postcode, parenting status). Usage indicators: frequency of visits, duration of participation over time, recency of last visit, number of new registrations, centre-level utilisation, repeat participation | Extracted by Local Authority data analyst from the Gladstone and 4global systems; cleaned and shared via data-sharing agreement |
| **Qualitative In-Depth Interviews (Service Users)** | Individuals who have accessed the Free-Swimming Programme in the past 12 months; paid swimmers who regularly swim at the centres | Experiences of using the programme, accessibility, perceived barriers/facilitators, cultural acceptability, impact on wellbeing and physical activity, comparison between FSP and paid sessions | Recruited via leisure centre staff, consent-to-contact lists within the LA leisure system, community outreach, PAG/PPI referrals, posters/newsletters at centres |
| **Qualitative In-Depth Interviews (Service Providers / Stakeholders)** | Leisure centre staff (frontline), swim coaches, public health staff, programme coordinators, senior managers | Implementation processes, contextual factors, resource and staffing implications, perceived outcomes, challenges and opportunities for sustainability and scale-up | Identified through stakeholder mapping; contacted via email through LA teams and leisure centre managers |
| **Focus Group Discussions (Non-Service Users)** | Eligible residents (women and girls 16+; men 55+) who have *not* engaged with the Free-Swimming Programme | Reasons for non-participation, awareness, perceived barriers, cultural and social considerations, accessibility, perceptions of leisure services | Recruitment via community boards/newsletters, local networks, leisure centres, PAG/PPI networks, snowball sampling |
| **Researcher Field Notes** | All qualitative participant groups | Contextual observations, reflections on interviews/FGDs, insights related to programme delivery and engagement | Generated by research team during/after interviews and FGDs |
| **Demographic Contextual Data** | Service users and non-users at population level | Borough demographic characteristics and population-level activity patterns | Local authority datasets, JSNA, Sport England Active Lives data |

### 4.6 Data Analysis

#### 4.6.1 Quantitative Data Analysis Plan

The quantitative component of the study will focus on identifying key factors associated with participation in swimming and perceived accessibility of leisure facilities among residents. Data will be entered, cleaned, and analysed using a statistical software package using R. The analysis will proceed in three main stages: descriptive analysis, bivariate analysis, and multivariate analysis.

##### Step 1: Descriptive Analysis

The first stage of analysis will summarise all variables to describe the study characteristics and identify outliers or missing data. Categorical variables will be presented using frequencies and percentages. Continuous variables (e.g., age in years, distance to the nearest leisure centre) will be summarised using appropriate measures of central tendency and spread. Age may also be grouped into age groups where helpful for interpretation. This stage will provide an overview of participant demographics, swimming participation patterns, and perceptions of accessibility, comfort, and affordability. Visual displays such as bar charts and histograms will be used to illustrate key patterns across subgroups.

##### Step 2: Bivariate Analysis

The second stage will explore relationships between the dependent and independent variables through bivariate statistical tests. The main dependent variable will be swimming participation, operationalised as frequency of swimming or engagement in weekly sessions (yes/no). Independent variables will include demographic, environmental, psychosocial, and service-related characteristics. Associations between categorical variables will be examined using Chi-square tests, while comparisons of continuous variables across groups will be assessed using independent t-tests or one-way ANOVA, depending on the number of comparison categories. For example, Chi-square tests will be used to examine whether swimming participation differs by gender. Similarly, t-tests or ANOVA will test for mean differences in perceived accessibility scores or comfort levels across income or age categories. Correlation analyses (Pearson or Spearman, depending on data normality) will also be conducted to assess relationships between continuous variables, such as distance to leisure centre and frequency of swimming.

##### Step 3: Multivariate Analysis

The third stage will employ multivariate regression models to identify independent predictors of swimming participation while adjusting for potential confounders. A binary logistic regression model will be fitted with weekly swimming participation (1 = yes, 0 = no) as the dependent variable. Key independent variables will include distance to the nearest leisure centre, awareness of free-swimming programmes, perceived confidence in swimming, household income, gender, and age. The model will estimate odds ratios (ORs) and 95% confidence intervals (CIs) to quantify the strength and direction of associations. Where participation frequency is measured on an ordinal scale (e.g., never, occasionally, weekly), ordinal logistic regression will be used instead. Where relevant, continuous predictors (e.g., age, distance) will be tested both as continuous and categorical variables to check for non-linear effects. All analyses will use a significance level of 0.05, and results will be reported with appropriate effect size measures and confidence intervals.

#### 4.6.2 Qualitative Data Analysis Plan

The qualitative data derived from focus group discussions, participatory sessions, and in-depth interviews will be analysed thematically using both deductive and inductive approaches. A deductive framework based on the RE-AIM model (Reach, Effectiveness, Acceptability, Implementation, and Maintenance) will provide an initial structure to guide coding and ensure that key dimensions of programme reach and impact are systematically explored. This framework will help identify how participants perceive accessibility, engagement, and the sustainability of swimming initiatives within their local context.

Alongside this, inductive coding will be employed to allow unanticipated themes and participant-driven insights to emerge organically from the data. Codes and themes will be iteratively refined through constant comparison, ensuring both theory-informed and data-grounded interpretations. Core themes are expected to cluster around concepts such as accessibility and comfort, cultural fit and inclusion, perceptions of value, and trust in service providers. NVivo analysis software be used to support the systematic management and organisation of codes and themes.

#### 4.6.3 Data Triangulation

To enhance the validity and credibility of the study findings, a triangulation strategy will be applied at both the data collection and analysis phases. During the data collection phase, triangulation will be achieved with multiple data sources and participant groups. This multi-perspective approach ensures that diverse experiences and contextual factors influencing swimming participation are captured, allowing for a richer and more nuanced understanding of the research problem. Different data collection tools, such as focus group discussions, in-depth interviews, participatory sessions, and administrative data from the Gladstone system, will be used to corroborate and complement each other.

At the analysis phase, triangulation will involve comparing and integrating findings across qualitative and quantitative strands to identify areas of convergence, divergence, and complementarity. Quantitative results such as demographic predictors of participation or facility usage trends will be interpreted alongside qualitative insights that explain the underlying motivations, perceptions, or barriers.

Furthermore, thematic patterns emerging from different participant groups will be cross-examined to assess consistency and contextual variation. The integration of findings will be guided by a mixed-methods logic of merging and connecting datasets, ensuring that the final interpretation provides a coherent, evidence-based understanding of swimming participation across demographic, environmental, and individual dimensions.

## 5.0 Ethics and governance

The study will seek ethical approval from the Loughborough University Ethics Committee.

### 5.1 Data Protection and Management

Only authorised members of the PHIRST Light research team and host institution staff responsible for monitoring or audit will have access to study data for regulatory compliance purposes. Data will be stored securely in the PHIRST Light OneDrive folder, hosted at the University of Nottingham and accessible to research members based at Loughborough University. Access to personal data will be restricted to the named study researchers and the Chief Investigator.

Research data and records will be retained for a maximum of seven years after study completion, after which they will be securely destroyed. In line with the Data Protection Act, personal data will be stored only for the study period and for no more than one year following submission of the final report to NIHR, as specified in the Participant Information Sheet (PIS) and consent form. Contact details provided by participants will be stored separately from the main dataset, accessible only to the research team.

A formal data-sharing agreement is being established between the Tower Hamlets Local Authority and the University of Nottingham. Under this agreement, a designated Local Authority data analyst will extract the relevant datasets required for the evaluation directly from Gladstone and 4global and undertake initial cleaning and preparation. This process will involve standardising and validating key variables to ensure consistency with the project’s data analysis plan and indicator framework. For example, aligning demographic categories such as ethnicity with national reporting standards. The cleaned and anonymised dataset will then be securely transferred as a CSV file to the research team using the University’s encrypted data transfer system for further analysis.

All data handling will comply with the General Data Protection Regulation (GDPR) and institutional data governance policies. The dataset shared with the research team will not contain any personally identifiable information. Data will be stored on encrypted, access-controlled university servers and used solely for the purposes of this evaluation. Only authorised members of the research team will have access, and all analyses will be conducted within secure university environments.

### 5.2 Risks

We recognise that this research may entail certain risks. Following careful consideration, we have outlined these potential risks below, along with strategies for how they can be appropriately managed and mitigated

We acknowledge that there are some risks associated with this research. Below, these risks have been outlined, with suggestions for how they might be mitigated/managed.

| Risks Identified | Mitigation Strategies |
| --- | --- |
| The structure or completeness of demographic and participation data may limit analysis. | So, there have early conversations with the LA to clarify data points and supplement missing data points with other administrative records. |
| There could be concerns with personal safety, including verbal abuse, assault, or unpredictable behaviour from members of the public towards researchers during in-person interviews | Physical interviews will be conducted in public venues.<br>Researchers keep charged phones and inform supervisors of arrival/departure.<br>Avoid visits after dark or outside normal working hours.<br>Pre-agree exit strategies and leave immediately if uncomfortable or threatened.<br>Never visit unplanned locations and avoid compromising situations.<br>Follow Loughborough University incident-reporting procedures. |
| Travelling to and from the interviews | Drive carefully and safely observing British highway law.<br>Only use licensed taxis, travel with people you know.<br>Public transport during working hours will be used where possible.<br>Hotel will be booked if during antisocial hours. |
| Participants may disclose mental health difficulties or safeguarding concerns during interviews | Interviewers will be trained in mental health first aid and have experience supporting individuals with mental health challenges. Any safeguarding concerns will be escalated promptly to the appropriate personnel (e.g., supervisors, line managers, or leisure centre managers) who are responsible for managing and responding to such issues. |
| Data storage of in-person, phone and online audio recordings | Researchers are instructed to observe data protection guidelines and to transfer the recordings from the audio recording equipment, and to store the data on the University's OneDrive as |
|  | soon as possible and to delete the recordings from the audio recording equipment |

## 6.0 Outputs

### 6.1 Dissemination plan

Study outcomes will be reported in a highlight project report and desk slides as requested by the Local Authority for the funder and service providers. We will actively engage with the local PPI group, PAG and wider stakeholder group to consider how the evaluation findings are most effectively communicated, alongside formalising a knowledge mobilisation plan. A lay summary will be made available for Tower Hamlets residents which will be circulated by the local authority. We will disseminate study findings through written paper(s) in academic journals, conferences and other stakeholder engagement and PPI events.

Broadly, dissemination will occur through the following channels:

- The PHIRST website, including publication of this protocol.
- Public facing summaries of the findings (print and web formats)
- Conference presentations and peer-reviewed, open access journal articles
- Creative outputs such as video, interactive content, or community events
- PHIRST Light and PHIRST social media channels
- Dissemination through professional networks
- Local Authority workshops and events

## Data Availability

All data produced in the present study will be available upon reasonable request to the authors

## LIST OF ABBREVIATIONS

A&E: Accident and Emergency
CMO: Chief Medical Officer
CSV: Comma-Separated Values
FSP: Free-Swimming Programme
FGD / FGDs: Focus Group Discussion(s)
GDPR: General Data Protection Regulation
HDRC: Health Determinants Research Collaboration
IDIs: In-Depth Interviews
JSNA: Joint Strategic Needs Assessment
LA: Local Authority
LBTH: London Borough of Tower Hamlets
NHS: National Health Service
NIHR: National Institute for Health and Care Research
PAG: Public Advisory Group
PIS: Participant Information Sheet
PPI: Patient and Public Involvement
PHIRST: Public Health Intervention Responsive Studies Team
RE-AIM: Reach, Effectiveness, Acceptability^*^, Implementation, Maintenance
UK: United Kingdom

## Footnotes

* This evaluation is adapting the RE-AIM framework to include acceptability instead of adoption. This adaptation is appropriate because acceptability focuses on how participants and providers experience, perceive, and respond to the programme in real-world contexts. This offers richer insight into the programme’s perceived value and suitability. In contrast, adoption examines differences between settings in terms of their capacity, resources, expertise, and willingness to implement the intervention.

## References

British Heart Foundation. (2017). New report assesses impact of physical inactivity on UK heart health and economy. https://www.bhf.org.uk/what-we-do/news-from-the-bhf/news-archive/2017/april/new-report-assesses-impact-of-physical-inactivity-on-uk-heart-health-and-economy

Eade, J. (2024). Bangladeshis in London and Tower Hamlets: Community activism and the local state. In Europe in a Global Context (pp. 25–47). 10.1007/978-3-031-53773-8_2

Green, F. (2017). British values and identity among young British Muslims in Tower Hamlets: Understandings and connections. Journal of Children’s Services, 12(4), 239–256. 10.1108/jcs-01-2017-0001

McKay, A., Tang, A., Dougall, D., Rumble, C., & Banerjee, S. (2014). Assessment of liver disease in Tower Hamlets, London, UK: Foundation for integrated solutions. The Lancet, 384, S50. 10.1016/S0140-6736(14)62176-0

Massey, H., Gorczynski, P., Harper, C. M., Sansom, L., McEwan, K., Yankouskaya, A., & Denton, H. (2022). Perceived impact of outdoor swimming on health: Web-based survey. Interactive Journal of Medical Research, 11(1), e25589. 10.2196/25589

Public Health England. (2016). Health matters: Getting every adult active every day. https://www.gov.uk/government/publications/health-matters-getting-every-adult-active-every-day/health-matters-getting-every-adult-active-every-day

Quin, M., Carmichael, L., & Hopper, C. (2022). Implementing Health Impact Assessment policy on infrastructure development in the London Borough of Tower Hamlets. Cities & Health, 1–9. 10.1080/23748834.2022.2148843

Shilton, T., Bauman, A., Beger, B., Chalkley, A., Champagne, B., Elings-Pers, M., Giles-Corti, B., Goenka, S., Miller, M., Milton, K., Oyeyemi, A., Ross, R., Sallis, J. F., Armstrong-Walenczak, K., Salmon, J., & Whitsel, L. P. (2024). More people, more active, more often for heart health – Taking action on physical activity. Global Heart, 19(1), 42. 10.5334/gh.1308

Smith, D., Kirkwood, G., Pott, J., Kourita, L., Jessop, V., & Pollock, A. M. (2015). Childhood injury in Tower Hamlets: Audit of children presenting with injury to an inner city A&E department in London. Injury, 46(6), 1131–1136. 10.1016/j.injury.2014.12.029

Sport England. (2025). Active Lives Adult Survey November 2023–24 report (pp. 4–11). https://sportengland-production-files.s3.eu-west-2.amazonaws.com/s3fs-public/2025-04/ActiveLivesAdult-Nov23-24_V9-23-04-25-10-03-03-02.pdf

Staley, K., Donaldson, A., Mosler, A. B., Seal, E., Forsyth, A., O’Halloran, P., Nicholson, M., Forsdike, K., & Bruder, A. M. (2024). The multi-dimensional impacts of injury on physically inactive women’s participation in sport and physical activity: Insights from concept mapping. Journal of Science and Medicine in Sport, 27(10), 716–725. 10.1016/j.jsams.2024.06.008

Swim England. (2017, June 25). Health and wellbeing benefits of swimming report. https://www.swimming.org/swimengland/health-and-wellbeing-benefits-of-swimming/

Tatari, E., & Teymuroglu, Z. (2023). A network analysis of minority representation in the British local government: Tower Hamlets Council (2006–2010). European Journal of Economic and Political Studies, 6(2), 19–35.

Tower Hamlets Council. (2025). Over 20,000 Tower Hamlets residents access free swimming. https://www.towerhamlets.gov.uk/News_events/2025/July/Over-20000-Tower-Hamlets-residents-access-free-swimmi cng.aspx

Tower Hamlets Public Health Department. (2016). Joint Strategic Needs Assessment summary document: Life, health, and wellbeing in Tower Hamlets. https://www.towerhamlets.gov.uk/Documents/Public-Health/JSNA/JSNA_Summary.pdf

Tower Hamlets Public Health Department. (2022). Tower Hamlets annual public health report (pp. 17–60). https://www.towerhamlets.gov.uk/Documents/Public-Health/TowerHamletsPublicHealthReport2022.pdf

Trust for London. (2025). Overview of London boroughs. https://tfl.ams3.cdn.digitaloceanspaces.com/media/documents/Tower_Hamlets_U0nD08j.pdf

